# Real-world experience on the use of nivolumab monotherapy for advanced renal cell carcinoma: a multicenter retrospective cohort study in Spain

**DOI:** 10.1101/2025.02.07.25321775

**Authors:** Natalia Fernández-Díaz, María Mateos-González, Ana Pertejo-Fernández, Juan Diego Cacho-Lavín, María José Juan-Fita, Isabel Chirivella-González, Mikel Arruti-Ibarbia, Ovidio Fernández-Calvo, Natalia Fernández-Núñez, María José Méndez-Vidal, Martín Lázaro-Quintela, Aurea Molina-Díaz, Nieves Del Pozo-Alonso, Olatz Etxaniz-Ulazia, Silvia Margarita García-Acuña, Yoel Z. Betancor, Ainara Azueta-Etxebarria, Ana Calatrava-Fons, Helena Lombardía-Rodríguez, Lorena Alarcón-Molero, Leire Etxegarai-Ganboa, Abraham Antón-Cameselle, José Antonio Bello-Giz, Carlos Manuel Neira-De Paz, Teresa Cabaleiro, Teresa González-Serrano, José Antonio Ortiz-Rey, Felipe Sacristán-Lista, Silvia García-Rubín, Elisa Ortega, Cristina Carrato-Moñino, Santiago Aguín-Losada, Luis León-Mateos, Jorge García-González, Álvaro Pinto-Marín, Ignacio Duran, Rafael López-López, Urbano Anido-Herranz, Juan Ruiz-Bañobre

## Abstract

The safety and efficacy of nivolumab in treating advanced renal cell carcinoma (aRCC) patients have been evaluated in real-world evidence (RWE) studies across various European countries, but no such data from Spain have been reported until now. We conducted a multicenter, retrospective study on 222 previously treated aRCC patients to assess the efficacy of nivolumab and the impact of pre-treatment factors on therapy outcomes in a real-world clinical practice scenario in Spain. Our results were then compared with other significant clinical experiences involving aRCC patients treated with nivolumab monotherapy. With a median follow-up of 14.6 months, the median overall survival (OS) was 18.1 months (95% confidence interval [CI], 14.2-23.7 months), and the median progression-free survival (PFS) was 4.96 months (95% CI, 3.98-7.13 months). The disease control rate was 51% (95% CI, 45-58%) and the objective response rate 23% (95% CI, 18-30%). Poor IMDC risk score was independently associated with worse OS, while prior nephrectomy with better OS. Poor IMDC risk score and ≥3 metastatic sites were independently associated with worse PFS; ≥3 metastatic sites was also associated with lower disease control. Consistent with prior clinical trials and RWE studies, this research confirms the efficacy and safety of nivolumab in daily clinical practice for a cohort of unselected previously treated aRCC patients in Spain.

## Introduction

Over the past few decades, the treatment of advanced renal cell carcinoma (aRCC) has evolved significantly. Initially, cytokine therapies such as interleukin-2 and interferon-alpha were standard, but their high toxicity limited their use. In the mid-2000s, targeted therapies emerged, starting with tyrosine kinase inhibitors sorafenib and sunitinib, which inhibit angiogenesis by blocking the VEGF pathway. These treatments led to substantial improvements in patient outcomes and were widely adopted as the standard of care for several years. However, most patients with aRCC eventually developed resistance to these therapies, leading to disease progression. Subsequently, mTOR inhibitors such as everolimus were introduced for patients who progressed on tyrosine kinase inhibitors^1-3^.

Fortunately, in November 2015, the U.S. FDA approved nivolumab (an anti-PD-1 antibody) for aRCC patients who had previously received treatment with anti-angiogenic agents^4,5^. A few months later, in April 2016, the European Medicines Agency (EMA) followed suit^6^. This approval, which initiated a revolution in the treatment landscape of aRCC, was based on the results of the multicenter, randomized, two-arm, open-label phase 3 CheckMate-025 (CM-025) trial, where nivolumab demonstrated a significant improvement in overall survival (OS) compared to everolimus, yielding a 27% reduction in the risk of death and an unprecedented median OS of 25 months^4^.

However, the results obtained in clinical trials, because of they are conducted in a rigorously controlled patient population, may not fully represent the real-world effectiveness of treatments when applied to an unselected general population with more variable clinical conditions^7^. Accordingly, we conducted this multicenter retrospective study in a cohort of previously treated aRCC patients to evaluate the efficacy of nivolumab and the influence of various pre-treatment factors on therapy outcomes in a real-world clinical practice scenario in Spain. Additionally, we compared our study results with the most relevant clinical experiences reported to date in similar scenarios involving aRCC patients undergoing nivolumab monotherapy.

## Patients and Methods

### Study design and patient population

We conducted a multicenter retrospective study involving a cohort of 222 patients with aRCC treated with nivolumab in either the second line of therapy or beyond in the context of routine clinical practice between December 2015 and October 2021 in thirteen Spanish medical centers (**Supplementary Table 1**). Patients received nivolumab at doses of either 3 mg/kg every 2 weeks, 240 mg every 2 weeks, or 480 mg every 4 weeks intravenously until disease progression or unacceptable toxicity.

**Table 1.**
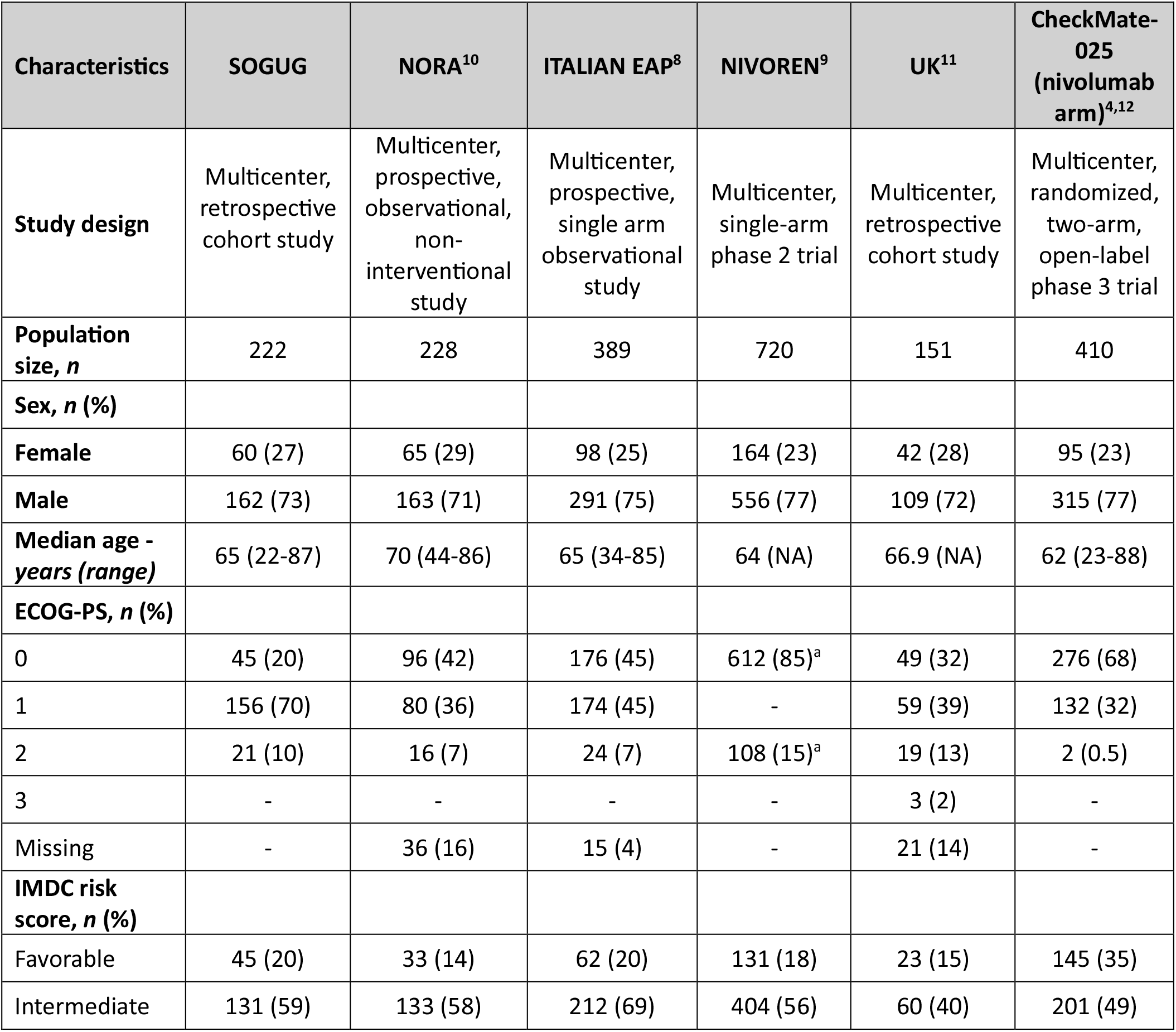
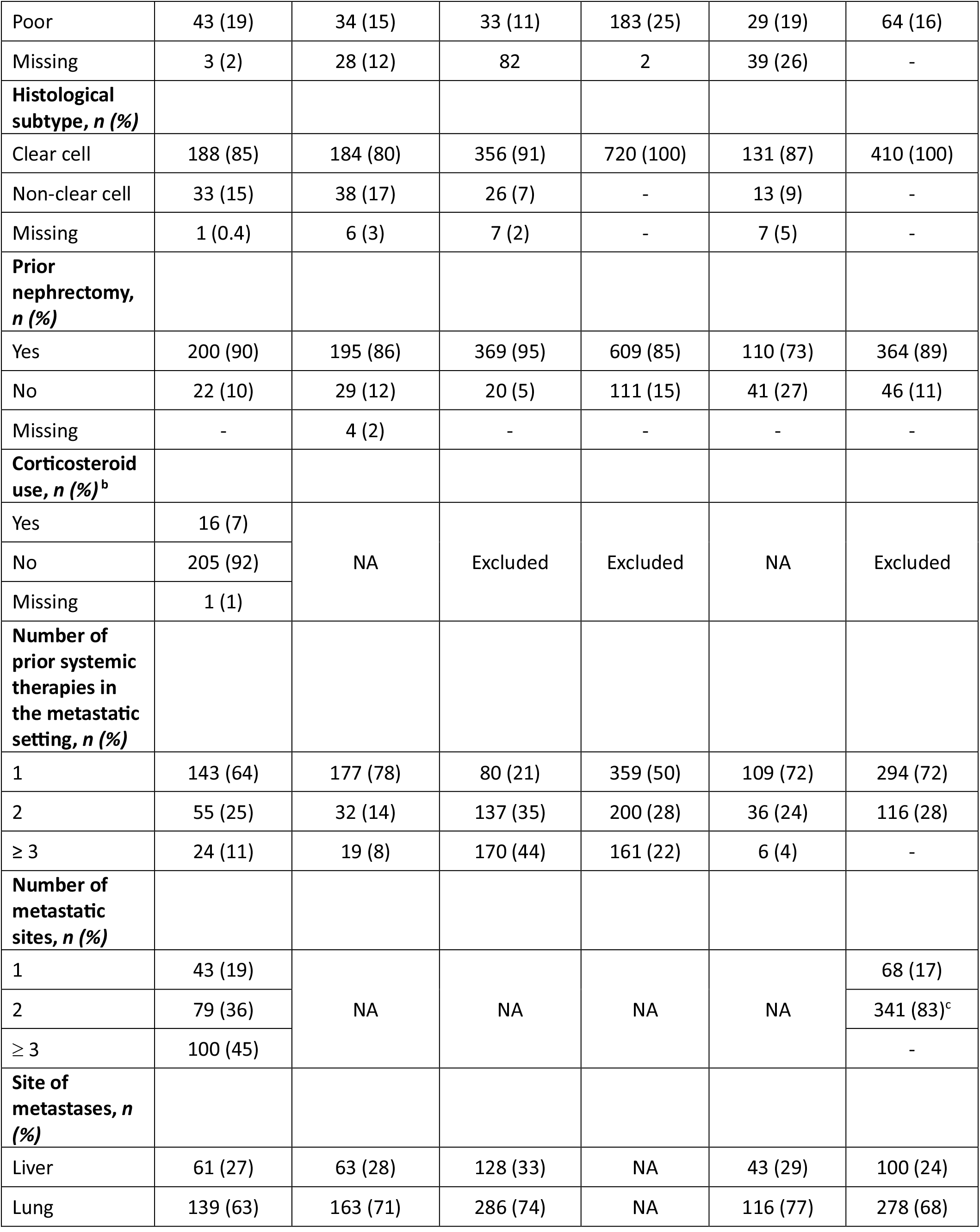
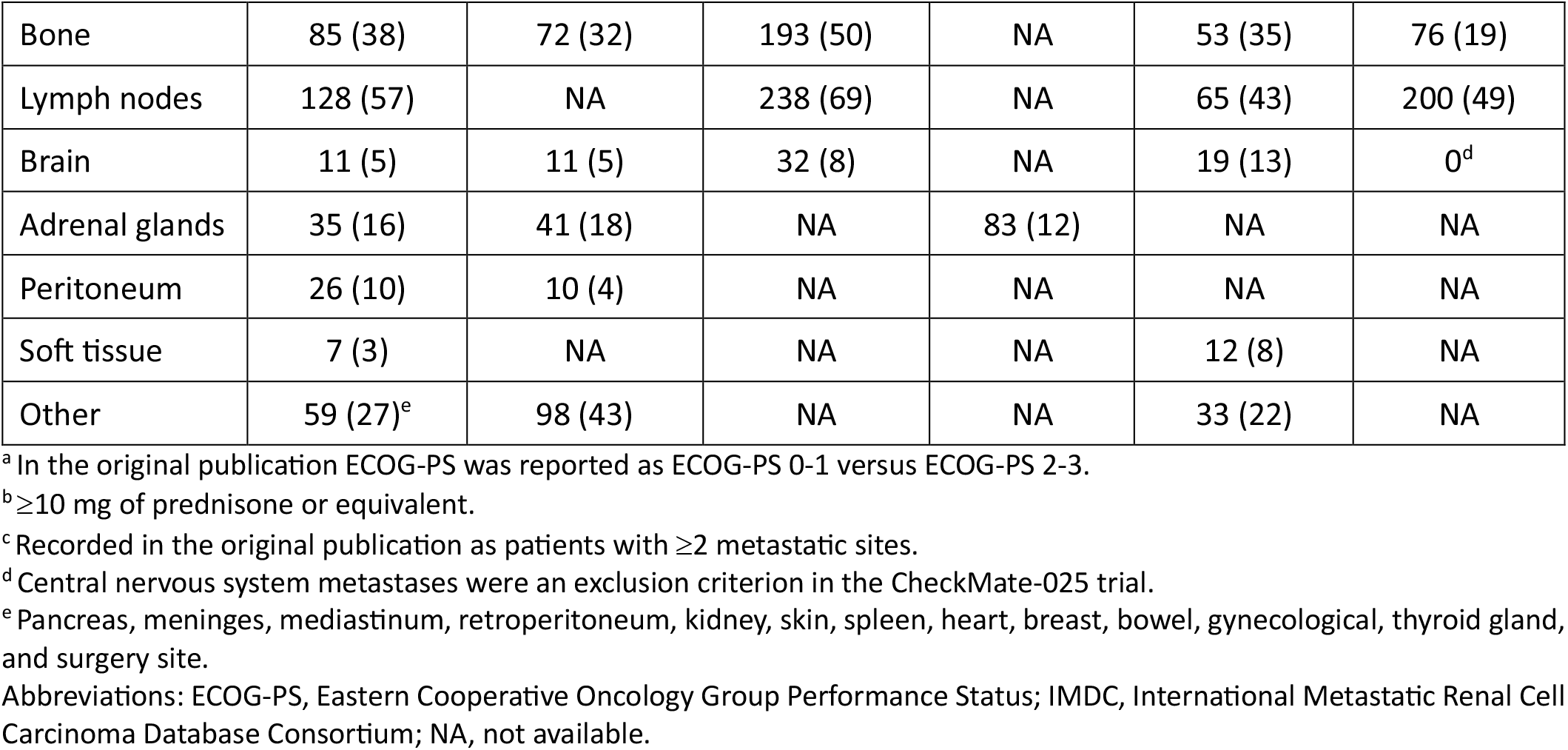
Baseline patient and disease characteristics for cases treated with nivolumab across different clinical trials and real-world evidence experiences.

Demographic, clinical, analytical and pathological data, as well as the use of corticosteroids were extracted form electronic medical records. We considered laboratory parameters, and corticosteroids use within a window of 30 days before the start of first nivolumab infusion.

The primary efficacy endpoint was OS. Secondary endpoints were progression-free survival, disease control rate (DCR), and overall response rate (ORR). Tumor responses were assessed following the local protocols of each center.

This study was approved by the Ethics Committee for Research with medicinal products of Galicia (2020/248), the Ethics Committee for Research with medicinal products of Euskadi (EPA2020065), the Ethics Committee for Research with medicinal products of Cantabria, the Ethics Committee for Research with medicinal products of Hospital Universitario La Paz (PI-4313), the Ethics Committee for Research with medicinal products of Córdoba (1584-N-20), and the Ethics Committee for Research at Hospital Universitario Elche-Vinalopó. The study was conducted in accordance with the guidelines of Good Clinical Practice and the Declaration of Helsinki. All living patients provided written informed consent before enrollment. Informed consent was waived for deceased patients before study initiation.

### Statistical analysis

OS was calculated from the date of nivolumab initiation until death resulting from any cause or last known follow-up for living patients. PFS was calculated from the date of nivolumab initiation until disease progression or death resulting from any cause or last known follow-up for patients with no disease progression. DCR was defined as the proportion of patients who achieved a complete or partial response and stable disease, and ORR as the proportion of patients who achieved a complete or partial response. Patients who died before radiologic assessment were considered not evaluable for response. Survival estimates were calculated by the Kaplan–Meier method, and groups were compared with the log-rank test. The Cox proportional hazards regression model was used to evaluate factors independently associated with OS and PFS. Baseline clinicopathological variables included in the multivariable analysis were selected according to statistical significance in univariable analysis (cutoff, P < 0.05). The proportional hazard assumption was verified with the Schoenfeld residual method. Factors associated with disease control (DC) and response were tested with logistic regression in univariable analyses. Variables included in the final multivariable model were selected according to their statistical significance in univariable analysis (cutoff, P < 0.05). All P-values were two-sided, and those less than 0.05 were considered statistically significant. All statistical analyses were performed using R version 4.3.2 (Vienna, Austria).

## Results

### Patient population

Between 10 December 2015 and 19 October 2021, a total of 222 patients were enrolled and received at least one dose of nivolumab. Baseline patient and disease characteristics are detailed in **Table 1**. The median age was 65 years (range, 22-87 years). Twenty-seven percent (n = 60) of patients were female, while 73% (n = 162) were male. Most patients (90%) had an Eastern Cooperative Oncology Group Performance Status (ECOG-PS) score of 0 (n = 45, 20%) or 1 (n = 156, 70%). Twenty percent (n = 45), 59% (n = 131), and 19% (n = 43) of patients had a favorable, intermediate and poor International Metastatic RCC Database Consortium (IMDC) risk score, respectively. Ninety percent (n = 200) of patients had undergone prior nephrectomy. Clear cell renal cell carcinoma (ccRCC) was the predominant histology, observed in 85% (n = 188) of patients. The most common metastatic sites were the lungs (n = 139, 63%), lymph nodes (n = 128, 57%), liver (n = 61, 27%), and bones (n = 85, 38%). Five percent (n = 11) of patients had central nervous system (CNS) metastases. **Table 1** compares the baseline patient and disease characteristics of our study with those of different clinical trials and real-world evidence experiences (RWE) studies on nivolumab in aRCC.

### Treatment exposure and safety

At the time of data collection, after a median follow-up of 14.6 months (range 0.1–88.4 months), the median number of nivolumab cycles administered was 7 (range 1–125). At that time, 91% of patients (n = 203) had discontinued treatment, with disease progression (70%, n = 156) being the most common cause for treatment discontinuation. Ten percent (n = 23) discontinued due to adverse events, 4% (n = 9) due to death, while 11% (n = 24) discontinued for other reasons. Treatment was ongoing in 9% (n = 19) of patients at the time of data collection. Nivolumab was used as second-line therapy in 64% of patients, as third-line therapy in 25%, and as fourth-line or later therapy in 11% of them. The most common therapies in the first-line setting were sunitinib (n = 120 patients, 54%) and pazopanib (n = 89 patients, 40%), while cabozantinib was the most common second-line therapy (n = 33 patients, 33%) (**Supplementary Table 2**).

**Table 2.** Efficacy endpoints across different clinical trials and real-world evidence experiences.

| Endpoints | SOGUG | NORA<sup>10</sup> | ITALIAN EAP<sup>8</sup> | NIVOREN<sup>9</sup> | UK<sup>11</sup> | CheckMate-025 (nivolumab arm)<sup>4,12</sup> |
| --- | --- | --- | --- | --- | --- | --- |
| Response, n (%) |  |  |  |  |  |  |
| Complete response | 15 (7) | 3 (1.3) | 3 (1) | 9 (1.3) | NA | 4 (1) |
| Partial response | 38 (17) | 43 (19) | 87 (22) | 135 (19.7) | NA | 99 (24) |
| Stable disease | 62 (28) | 60 (26) | 124 (32) | 214 (31.1) | NA | 141 (34) |
| Progressive disease | 93 (42) | 75 (33) | 141 (36) | 329 (47.9) | NA | 143 (35) |
| Not evaluable | 14 (6) | 47 (21) | 34 (9) | 7 | NA | 23 (6) |
| ORR % (95% CI) | 23 (18-30) | 20 | 23 | 20.8 | NA | 23 (19-27) |
| DCR % (95% CI) | 52 (45-58) | 46 | NA | NA | NA | NA |
| Median OS, months (95% CI) | 18.05 (14.2-23.7) | 24.3 (19-28) | NA | 24.5 | 19.2 (16.9-27) | 25.8 (22.2-29.8) |
| 6-month OS rate, % (95% CI) | 78 (73-84) | 79 | 80 (76-84) | NA | NA | NA |
| 12-month OS rate, % (95% CI) | 62 (56-69) | 65 (59-71) | 63 (58-68) | 69 (66-73) | NA | 76 |
| 24-month OS rate, % (95% CI) | 42 (36-49) | 51 (44-58) | NA | NA | NA | 52 |
| 36-month OS rate, % (95% CI) | 28 (22-35) | 37 (28-45) | NA | NA | NA | 39 |
| Median PFS, months (95% CI) | 4.96 (3.98-7.1) | 5.3 (3.9-6.7) | 4.4 (3.7-6.2) | 3.2 (2.9-4.6) | NA | 4.2 (3.7-5.4) |
| Median nivolumab doses (range) | 7 (1-125) | 10 (1-96) | 13 (1-49) | 10 (1-30) | NA | NA |

Abbreviations: CI, confidence interval; DCR, disease control rate; NA: Not available; NR, not reached; ORR, overall response rate; OS, overall survival; PFS, progression-free survival.

### Efficacy

**Overall Survival**. At the time of data collection, 73% (n = 163) of enrolled patients had died. Median OS was 18.1 months (95% CI, 14.2-23.7) (**Table 2, Figure 1A**) and the 6-, 12-, 24- and 36-month OS rates were 78% (95% CI, 73-84), 62% (95% CI, 73-84), 42% (95% CI, 36-49) and 28% (95% CI, 22-35), respectively (**Table 2**). Of ten baseline variables examined, four were associated with worse OS in univariable analysis: ECOG-PS ≥2 (HR = 2.5, 95% CI, 1.5-4.1; P = 0.0002), poor IMDC risk score (HR = 2.68, 95% CI, 1.86-3.89; P < 0.0001), >1 prior therapy for metastatic disease (HR = 1.4, 95% CI, 1-1.9; P = 0.047), and ≥3 metastatic sites (HR = 1.61, 95% CI, 1.2-2.2; P = 0.0025); and one with better OS: prior nephrectomy (HR 0.58, 95% CI, 0.35-0.94; P = 0.029). Of them, only two remained independently associated with OS in multivariable analysis: poor IMDC risk score (HR 1.97, 95% CI, 1.33-2.95; P = 0.0009) with worse OS, and prior nephrectomy (HR 0.58, 95% CI, 0.35-0.98; P = 0.04) with better OS (**Table 3**).

**Table 3.** Univariable and multivariable Cox regression analyses for overall survival and progression-free survival.

| Characteristics | Univariable analysis |  | Multivariable analysis |  |
| --- | --- | --- | --- | --- |
|  | HR (95% CI) | P value | HR (95% CI) | P value |
| **Overall survival** |  |  |  |  |
| Sex (Male vs female) | 1.12 (0.79-1.59) | 0.52 | - | - |
| Age (<75 vs ≥75 years) | 1.2 (0.85-1.8) | 0.26 | - | - |
| ECOG PS (2 vs 0-1) | 2.5 (1.5-4.1) | **0.0002** | 1.57 (0.91-2.71) | 0.1 |
| IMDC risk score (poor vs intermediate/good) | 2.68 (1.86-3.89) | **< 0.0001** | 2.06 (1.36-3.12) | **0.0007** |
| Histology (ccRCC vs non-ccRCC) | 0.83 (0.54-1.3) | 0.42 | - | - |
| Prior nephrectomy (yes vs no) | 0.58 (0.35-0.94) | **0.029** | 0.58 (0.35-0.98) | **0.04** |
| Corticosteroid use (yes vs no) | 1.03 (0.56-1.9) | 0.92 | - | - |
| CNS metastasis (yes vs no) | 0.74 (0.35-1.6) | 0.45 | - | - |
| Number of prior therapies (>1 vs 1) | 1.4 (1-1.9) | **0.047** | 1.34 (0.95-1.88) | 0.09 |
| Number of metastatic sites (≥3 vs < 3) | 1.6 (1.2-2.2) | **0.0025** | 1.32 (0.95-1.83) | 0.1 |
| **Progression-free survival** |  |  |  |  |
| Sex (Male vs female) | 1.02 (0.74-1.39) | 0.92 | - | - |
| Age (<75 vs ≥75 years) | 1.34 (0.94-1.92) | 0.11 | - | - |
| ECOG PS (2 vs 0-1) | 1.57 (0.97-2.52) | 0.06 |  |  |
| IMDC risk score (poor vs intermediate/good) | 1.83 (1.29-2.59) | **0.0007** | 1.69 (1.19-2.41) | **0.003** |
| Histology (ccRCC vs non-ccRCC) | 0.77 (0.52-1.14) | 0.2 | - | - |
| Prior nephrectomy (yes vs no) | 0.84 (0.52-1.34) | 0.46 | - | - |
| Corticosteroid use (yes vs no) | 1.27 (0.74-2.16) | 0.38 | - | - |
| CNS metastasis (yes vs no) | 0.89 (0.47-1.69) | 0.73 | - | - |
| Number of prior therapies (>1 vs 1) | 1.28 (0.95-1.71) | 0.1 | - | - |
| Number of metastatic sites (≥3 vs < 3) | 1.59 (1.20-2.11) | **0.0012** | 1.50 (1.12-1.99) | **0.006** |

Bold numbers indicate statistically significant values.
Abbreviations: ccRCC, clear cell renal cell carcinoma; CI, confidence interval; CNS, central nervous system; ECOG-PS, Eastern Cooperative Oncology Group Performance Status; IMDC, International Metastatic Renal Cell Carcinoma Database Consortium; HR, hazard ratio.

**Figure 1.**
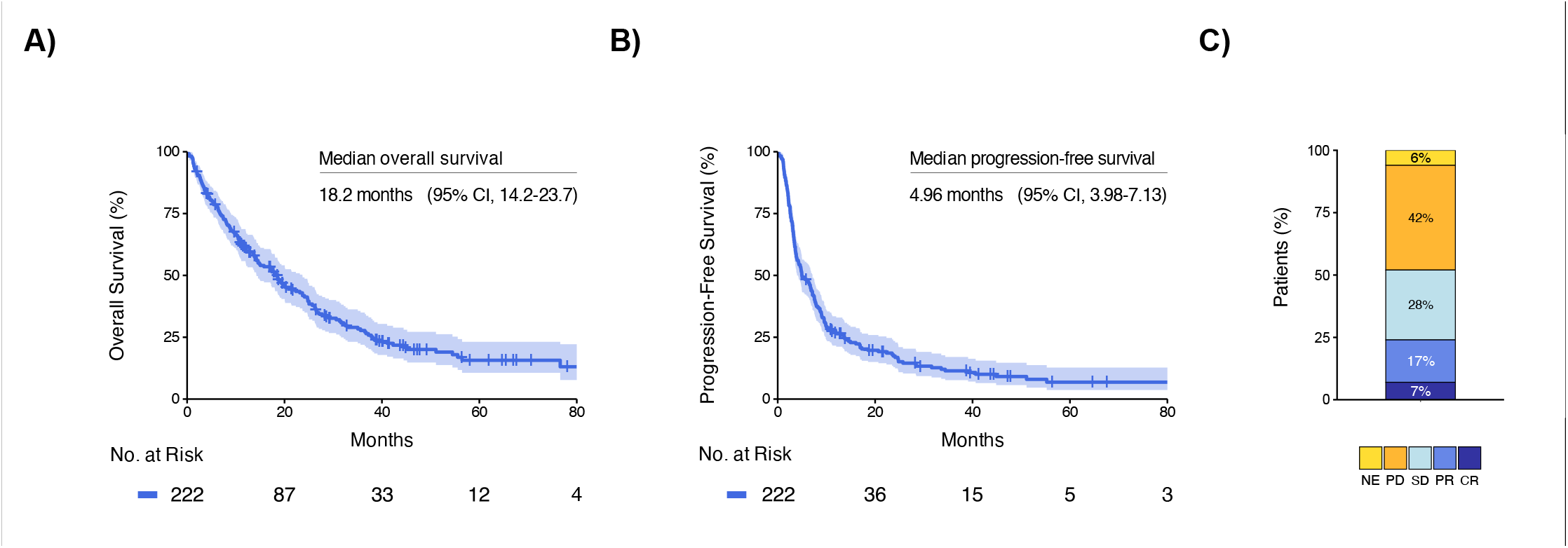
Efficacy of nivolumab in the overall population. **A**. Overall survival. **B**. Progression-free survival. **C**. Best response. Abbreviations: CR, complete response; NE, not evaluable; OS, overall survival; PD, progressive disease; PFS, progression-free survival; PR, partial response; SD, stable disease.

#### Progression-Free Survival

Median PFS was 4.96 months (95% CI, 3.98-7.13) (**Table 2, Figure 1B**). Of ten baseline variables examined, two were associated with worse PFS in univariable analysis: poor IMDC risk score (HR = 1.83, 95% CI, 1.29-2.59; P = 0.0007) and ≥3 metastatic sites (HR = 1.59, 95% CI, 1.20-2.11; P = 0.0012). Of them, both remained independently associated with worse PFS in multivariable analysis: poor IMDC risk score (HR = 1.69, 95% CI, 1.19-2.41; P = 0.003) and ≥3 metastatic sites (HR = 1.50, 95% CI, 1.12-1.99; P = 0.006) (**Table 3**).

#### Disease Control Rate and Objective Response Rate

DCR and ORR were 51% (95% CI, 45-58) and 23% (95% CI, 18-30), respectively, including 15 (7%) complete responses and 38 (17%) partial responses (**Table 2 and Figure 1C**). Of 10 baseline variables examined, none were associated in univariable analysis with lower ORR but 4 were associated with lower DCR: ECOG-PS ≥2 (OR = 0.34, 95% CI, 0.12-0.88; P = 0.03), poor IMDC risk score (OR = 0.38, 95% CI, 0.19-0.76; P = 0.008), > 1 prior therapies (OR = 0.51, 95% CI, 0.28-0.88; P = 0.02), and ≥3 metastatic sites (HR = 0.34, 95% CI, 0.20-0.59; P = 0.0001). Of them, only one remained independently associated with lower DCR: ≥3 metastatic sites (OR = 0.40, 95% CI, 0.23-0.70; P = 0.002) (**Supplementary Tables 3 and 4**).

## Discussion

The treatment landscape of aRCC has undergone significant transformation since the approval of nivolumab in November 2015 for patients with accRCC who received previous antiangiogenic therapy. This approval was based on the results of the multicenter, two-arm, randomized, open-label, phase 3 CM-025 clinical trial, which demonstrated that nivolumab not only improved OS and ORR compared to everolimus among accRCC patients but also a more favorable safety profile.

In recent years, RWE studies have gained substantial prominence within the clinical oncology community. This shift is due to the fact that clinical trials, conducted in highly controlled clinical settings, even though they are fundamental today, often do not fully recapitulate the diversity and complexity of patients encountered in real-world clinical practice. For instance, in the CM-025 clinical trial, patients with CNS metastases, prior mTOR inhibitor treatment, active/suspected autoimmune diseases, uncontrolled adrenal insufficiency, active chronic liver disease, and non-clear cell RCC were excluded. Furthermore, patients on systemic corticosteroids (>10 mg daily prednisone or equivalent) or other immunosuppressant agents were also excluded^4^.

Therefore, real-world studies are crucial for extending evidence on efficacy and safety to a broader patient population, more faithfully representing the daily clinical practice scenario^7.^ Examples of such studies in aRCC include the Italian early access program (EAP)^8^, the GETUG-AFU 26 NIVOREN trial^9^, the NORA study^10^, and the UK real-world experience^11^, which provide additional perspectives on the impact of nivolumab in real-world clinical settings across different European countries.

Considering this, we conducted a multicenter retrospective study involving a cohort of 222 previously treated aRCC patients to evaluate the efficacy of nivolumab and the influence of different baseline factors on therapy outcomes in a daily clinical practice scenario in Spain. Furthermore, we evaluated similar experiences published from other countries and compared our study results to theirs.

In our study, the real-world efficacy of nivolumab appeared comparable to that reported in previous experiences. The median OS was 18.05 months, which is lower than the 25.8 months observed in the CM-025^4,12,13^ and other RWE studies such as the NORA^10^ and NIVOREN^9^ trials, but similar to the Italian EAP^8^ and the UK real-world experience^11^. Moreover, the median PFS was 4.96 months, slightly higher than that reported in the CM-025 trial and other RWE experiences, except for the NORA study, which reported a median PFS of 5.3 months. Regarding therapy response, our study showed an ORR of 23%, which again was consistent with CM-025 trial^4,12^ and other RWE experiences. Notably, our study yielded a higher rate of complete responses (7%) compared to ∼1% in other studies^4,8-12^. However, our study also showed a greater proportion of progressive disease as the best response to nivolumab (42%), like the NIVOREN trial (48%)^9^.

Regarding the influence of different patient and disease characteristics on nivolumab outcomes, several aspects deserve further attention.

First, the median age of our study population was very similar to that reported in other studies^4,8-12^, ranging between the sixth and seventh decades. However, it is noteworthy that the CM-025 trial had a notably higher proportion of patients aged ≥75 years (18%) compared to our study (8%). Consistent with previous experiences, there was a predominance of men over women, with a ratio of 2.5-3:1, which slightly differs from the traditionally reported male/female incidence rate of 2:1. This discrepancy in sex distribution might be explained by the fact that the proportion of female patients presenting with aRCC appears to be significantly lower than that of males, as reported by other groups^14,15^. Importantly, we did not find statistically significant differences in terms of survival or response by age (<75 *vs* ≥75 years) or sex (male *vs* female).

Second, compared with some of the previous studies^4,10,11^, our population included more patients with less favorable baseline characteristics, such as those receiving three or more prior systemic therapies (11%) or those with a poor performance status (ECOG-PS 2, 10%). Regarding baseline characteristics associated with worse survival outcomes, although four variables were associated in univariable analyses with a higher risk of death (ECOG PS ≥2, poor IMDC risk score, >1 prior therapy for metastatic disease, and ≥3 metastatic sites), only poor IMDC risk score remained statistically significant after adjusting for various confounding factors in the multivariable Cox regression analysis. Interestingly, and consistent with a previous report by Rebuzzi et al^16^, nephrectomy prior to nivolumab initiation again emerged as a baseline feature independently associated with a significantly reduced risk of death. On the other hand, only poor IMDC risk score and having ≥3 metastatic sites were independently associated with worse PFS. Furthermore, having ≥3 metastatic sites was the only variable associated with disease control, while no baseline characteristics were correlated with response.

Third, unlike CM-025, our study included patients with CNS metastasis, non-clear cell renal cell carcinoma histology, and prior corticosteroid use at a dose of >10 mg/day of prednisone or its equivalent. Interestingly, none of these characteristics significantly impacted efficacy outcomes. In this regard, a cohort study from the NIVOREN trial suggests that untreated brain metastases may reduce the efficacy of immunotherapy, emphasizing the potential benefit of combining focal brain therapy with immune checkpoint inhibitors^17^.

This study presented some methodological limitations. First, it included data from 222 patients across thirteen medical centers. Although geographically dispersed within the Spanish healthcare system, this sample may not fully represent the broader aRCC patient population or treatment centers across Spain as a whole. On a positive note, this is the first and unique report of the use of nivolumab in this clinical scenario in Spain. Second, although adverse events were not recorded, we included the reasons for the discontinuation of nivolumab monotherapy. Third, the fact that tumor responses were assessed following the local protocols of each center could introduce some bias in the PFS assessment. Fortunately, this bias is mitigated by the availability of OS data and a wide follow-up period for patients alive at the end of the study period. Lastly, the use of retrospective medical record data could have introduced bias due to incomplete or inaccurate information in the original records, as well as potential errors in electronic case report form data entry. Nonetheless, to minimize these issues inherent to the retrospective nature of the study, built-in validation checks to the electronic case report form were applied.

This study, consistent with previous clinical trials and RWE experiences, confirms for the first time the efficacy and safety of nivolumab in daily clinical practice for a cohort of unselected, previously treated aRCC patients in Spain. Although the use of nivolumab monotherapy has decreased in recent years with the introduction of anti-PD-1-based combinations in the first line setting, it remains the treatment of choice for patients who, due to either access or prognostic reasons, have received tyrosine kinase inhibitor monotherapy as their initial treatment for metastatic disease. The granularity of the available data, as well as the accessibility of tumor samples from multiple cases in this cohort, warrants the future development of clinically relevant translational studies. These studies will aim to decipher the determinants of response to nivolumab and pave the way for a more rational and appropriate use of immunotherapy in aRCC patients.

## Acknowledgements

JR-B is supported by a Juan Rodés contract (JR21/00019) from the Institute of Health Carlos III.

## Author Contributions

Conceptualization: NF-D and JR-B; so|ware, NF-D and JR-B; validation, NF-D and JR-B; formal analysis, NF-D and JR-B; investigation: all authors; resources: all authors; data curation, all authors; writing – original dra|, NF-D and JR-B; writing – review & editing, all authors; visualization, NF-D, YZB, and JR-B; supervision, JR-B; project administration, JR-B; funding acquisition, JR-B.

## Data availability

The data that support the findings of this study are available from the corresponding author upon reasonable request.

