## Supplementary Material for "Real-world experience on the use of nivolumab monotherapy for advanced renal cell carcinoma: a multicenter retrospective cohort study in Spain"

Supplementary Tables:

- Supplementary Table 1
- Supplementary Table 2
- Supplementary Table 3
- Supplementary Table 4

**Supplementary Table 1**. Number of patients included per medical center.

| **Medical center** | **Patients, *n (%)*** |
|---|---|
| CHUAC | 11 (5) |
| CHUO | 14 (6) |
| CHUS | 49 (22) |
| CHUVI | 12 (5) |
| HCUV | 16 (7) |
| HGU | 16 (7) |
| HULA | 13 (6) |
| HUMV | 21 (10) |
| HULP | 26 (12) |
| HURS | 12 (5) |
| ICO | 5 (2) |
| IVO | 19 (9) |
| HUV | 8 (4) |

Abbreviations: CHUAC, Complejo Hospitalario Universitario de A Coruña; CHUO, Complejo Hospitalario Universitario de Ourense; CHUS, Complejo Hospitalario Universitario de Santiago de Compostela; CHUVI, Complejo Hospitalario Universitario de Vigo; HCUV, Hospital Clínico Universitario de Valencia; HGU, Hospital Galdakao-Usansolo; HULA, Hospital Universitario Lucus Augusti; HUMV, Hospital Universitario Marqués de Valdecilla; HULP, Hospital Universitario La Paz; HURS, Hospital Universitario Reina Sofía; ICO, Catalan Institute of Oncology; IVO, Instituto Valenciano de Oncología; HUV, Hospital Universitario del Vinalopó.

**Supplementary Table 2.** Drugs received prior to nivolumab.

| Characteristics | Patients |
|---|---|
| **First line, *n (%)*** | 222 |
| **Pazopanib** | 89 (40) |
| **Sunitinib** | 120 (54) |
| **Other** | 13 (6) |
| **Second line, *n (%)*** | 79 |
| Axitinib | 18 (26) |
| Cabozantinib | 23 (33) |
| Everolimus | 13 (19) |
| Pazopanib | 9 (13) |
| Sunitinib | 7 (11) |
| Other | 9 (13) |
| **Third line or beyond, *n (%)*** | 24 |
| Axitinib | 11 |
| Cabozantinib | 7 |
| Everolimus | 5 |
| Sunitinib | 4 |
| Other | 8 |
| **Median nivolumab doses, *n (range)*** | 7 (1-125) |
| **Discontinued treatment, *n (%)*** | 203 (91) |
| **Reasons for discontinuation, *n (%)*** | |
| Progression | 156 (70) |
| Death | 9 (4) |
| Adverse event | 23 (10) |
| Other | 24 (11) |

**Supplementary Table 3.** Univariable logistic regression analyses for response.

| Characteristics | Univariable analysis | |
|---|---|---|
| | **OR (95% CI)** | **P value** |
| Sex (male *vs* female) | 0.86 (0.45-1.81) | 0.74 |
| Age (<75 *vs* ≥75 years) | 0.54 (0.19-1.29) | 0.2 |
| ECOG PS (≥2 *vs* 0-1) | 0.32 (0.05-1.15) | 0.13 |
| IMDC risk score (poor *vs* intermediate/good) | 0.49 (0.18-1.16) | 0.13 |
| Histology (ccRCC *vs* non-ccRCC) | 1.13 (0.48-2.99) | 0.78 |
| Prior nephrectomy (yes *vs* no) | 0.80 (0.31-2.33) | 0.65 |
| Corticosteroid use (yes *vs* no) | 0.76 (0.17-2.48) | 0.68 |
| CNS metastasis (yes *vs* no) | 1.94 (0.49-6.71) | 0.31 |
| Number of prior therapies (>1 *vs* 1) | 0.53 (0.25-1.03) | 0.07 |
| Number of metastatic sites (≥3 *vs* < 3) | 0.70 (0.37-1.32) | 0.28 |

Abbreviations: ccRCC, clear cell renal cell carcinoma; CI, confidence interval; ECOG-PS, Eastern Cooperative Oncology Group Performance Status; IMDC, International Metastatic Renal Cell Carcinoma Database Consortium; OR, odds-ratio.

**Supplementary Table 4.** Univariable and multivariable logistic regression analyses for disease control.

| Characteristics | Univariable analysis | | Multivariable analysis | |
|---|---|---|---|---|
| | OR (95% CI) | P value | OR (95% CI) | P value |
| Sex (male *vs* female) | 0.90 (0.49-1.62) | 0.72 | - | - |
| Age (<75 *vs* ≥75 years) | 0.53 (0.26-1.07) | 0.08 | - | - |
| ECOG PS (≥2 *vs* 0-1) | 0.34 (0.12-0.88) | **0.03** | 0.49 (0.16-1.39) | 0.19 |
| IMDC risk score (poor *vs* intermediate/good) | 0.38 (0.19-0.76) | **0.008** | 0.57 (0.26-1.24) | 0.16 |
| Histology (ccRCC *vs* non-ccRCC) | 1.51 (0.72-3.24) | 0.28 | - | - |
| Prior nephrectomy (yes *vs* no) | 1.97 (0.81-5.14) | 0.14 | - | - |
| Corticosteroid use (yes *vs* no) | 0.94 (0.33-2.65) | 0.91 | - | - |
| CNS metastasis (yes *vs* no) | 2.64 (0.74-12.31) | 0.16 | - | - |
| Number of prior therapies (>1 *vs* 1) | 0.51 (0.28-0.88) | **0.02** | 0.58 (0.32-1.05) | 0.07 |
| Number of metastatic sites (≥3 *vs* < 3) | 0.34 (0.20-0.59) | **0.0001** | 0.40 (0.23-0.70) | **0.002** |

Bold numbers indicate statistically significant values.

Abbreviations: ccRCC, clear cell renal cell carcinoma; CI, confidence interval ECOG-PS, Eastern Cooperative Oncology Group Performance Status; IMDC, International Metastatic Renal Cell Carcinoma Database Consortium; OR, odds-ratio.
